# Statistical Methods for Estimating Cure Fraction of COVID-19 Patients In India

**DOI:** 10.1101/2020.05.30.20117804

**Authors:** E. P. Sreedevi, P. G. Sankaran

## Abstract

The human race is under the COVID-19 pandemic menace since beginning of the year 2020. Even though the disease is easily transmissible, a massive fraction of the affected people are recovering. Most of the recovered patients will not experience death due to COVID-19, even if they observed for a long period. They can be treated as long term survivors (cured population) in the context of lifetime data analysis. In this article, we present some statistical methods to estimate the cure fraction of the COVID-19 patients in India. Proportional hazards mixture cure model is used to estimate the cure fraction and the effect of covariates gender and age on lifetime. The data available on website ‘https://api.cvoid19india.org’ is used in this study. We can see that, the cure fraction of the COVID-19 patients in India is more than 90%, which is indeed an optimistic information.

## 1. Introduction

The outbreak of novel coronavirus disease 2019 (COVID-19) has created a global health crisis since January 2020. The first case of COVID-19 was reported in Wuhan, Hube Province, China as a case of pneumonia of unknown etiology on 31 December 2019. By May 19,2020 the virus has spread all over the world. It affected nearly 50 lakhs (4982937) people and caused more than three lakhs and twenty thousand (324554) deaths while more than 19 lakhs people(1958416) people recovered from the disease. The World Health Organization (WHO) declared the novel coronavirus (COVID-19) outbreak a global pandemic on March 11, 2020. However, in India the first case of COVID-19 is reported in the state of Kerala on 30 January 2020, for a medical student came from China. Now, all major cities in India are under the threat of COVID-19. By this time, 106480 positive cases were reported and death toll reached 3301. The good news is that a large group of people, counted to be 42309 recovered from the disease. The development a of medical facilities and the rapid actions in taking treatments, make sure that a lion’s share of the patients are recovered completely from the disease.

The popular statistical models in the analysis of epidemiological data is SIR (Susceptible, Infected and Recovered) model and allied other compartmental models. Many studies were reported on COVID-19 patient data using SIR models and other popular statistical techniques (Nadia and Hazem, 2020 and Waquas et al, 2020). In the present paper, our goal is to analyse COVID-19 patient data from India, using lifetime data models which is widely used in epidemiological studies and public health research (Lee and Go, 1997 and Cole and Hudgens, 2010).

In lifetime data analysis we are modelling time to occurrence of an event of interest. In this context, we can define death due to COVID-19 as the event of interest. The time is known as lifetime. Due to various reasons, the final occurrence time of the event is not available for many individuals (patients). This situation leads to the phenomena known as censoring. For censored patients we are only available with the partial information on lifetime; it is greater than a particular time. Possibility of censoring makes lifetime data analysis differ from all other fields of statistics. An extensive review of lifetime data analysis is given in Lawless (2011). For many COVID-19 patients in India, date of conformation of the disease and date of death/ recovery due to (from) the disease along with information on age and gender are available. The data is freely accessible in the site ‘https://api.cvoid19india.org’. When the event is defined as death due to COVID-19, the recovered and hospitalized patients can be treated to have censored lifetimes, since we do not have information on them after the given date (as per records).

In lifetime studies, researchers generally assume that all of the study subjects will experience the event of interest, if they are followed long enough (Maller and Zhao, 1996). However, in some situations a non-negligible proportion of individuals may not experience the event of interest even after a long period of time. For example a COVID-19 patient recovered once from the disease is assumed that he/she has acquired immunity to the disease and will not experience it in future. These patients can be treated as long term survivors or cured patients. Cure models are treated as an effective statistical tool for analysing data epidemiological studies and clinical cancer research (Othus et al., 2012 and Stolenberg et al., 2020).

Accordingly, the COVID-19 affected population in India includes many long term survivors, and an estimate of the cure fraction alludes to an estimate of the percentage of individuals who will recover. We should note that censored individuals and cured individuals are absolutely different; where the former case referred to an individual who does not experience the event of interest at a given time (censoring time) while the latter case referred to an individual who will not experience the event of interest even if they observed infinitely long. We can use mixture cure models, the basic cure model proposed by Boag (1949) to analyse this data. The mixture cure model assumes that the population under study is a mixture of susceptible (uncured)individuals who may experience the event of interest, and non-susceptible (cured) individuals who will never experience the event. Cox’s PH model (Cox, 1972) for cured data is used to investigate the association between survival time and covariates.

In the present paper, we analyse the data on COVID-19 patients in India (available as on 19 May, 2020) using statistical techniques in lifetime data analysis. This appears to be the first study on COVID-19 patient data in this direction. The rest of the article is organised as follows. In Section 2, we describe the statistical models and computational procedures employed in this study. We use Kaplan Meier estimate of the survivor function to get a basic information about the presence of long term survivors in the population. The effect of covariates on lifetime is analysed using proportional hazards model and the cure fraction is estimated in presence of covariates. Here the regression parameters as well as the cure fraction are estimated simultaneously. We use the package ‘smcure’ in R language (Cai et al., 2012) for this purpose. In Section 3, we present the results and analyse them with a detailed discussion. Finally, Section 4 gives concluding remarks and possible future works.

## 2. Model and Inference Procedures

### 2.1. Model

In general, suppose that we have n patients under study. Define *T* as the time to death of a COVID-19 patient from the date of confirmation of the disease. Hence *T* will be counted as number of days in hospital. The number of hospitalised days for patient whose death is happened is counted as an observed lifetime. If the patient is recovered we know only that presently the event is not happened to the patient, hence the hospitalised number of days for those patients are considered as censored lifetime. For patients with current status as ‘hospitalized’ also, no information is available after the given date. Hence those lifetimes are also treated as censored lifetimes.

The main frame works of lifetime data analysis are the survivor function, hazard rate function and distribution function. The survivor function denoted by *S*(*t*), gives the probability of a patient surviving beyond a time *t* and is defined by *S*(*t*) = *P*(*T > t)*. In general, we have survivor function is a non-increasing function with lim*_t_*_→∞_ *S*(*t*) = 0. The hazard rate function represented by *h(t)* indicates the instantaneous probability of failure at time *t* and it is given by 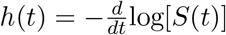. Distribution function defined as the complement of survivor function, gives the probability of failure before a particular time *t*, and is defined as *F*(*t*) = *P*(*T ≤ t*) = 1 − *S*(*t*).

The information on age and gender are available for some patients. The covariates can be represented using vector *z*, which is missing partially or completely for some patients. In general, we assume the vector *z* has *p ×* 1 dimension. Now for each patient, we observe a lifetime *T* and an indicator variable *δ*, which is defined as 0 if death is observed and 1 if the patient is recovered/hospitalized. Now, each patient has a pair of observation {*T*, *δ*}, the observed lifetime and an indicator variable to tell us whether *T* is an event time or censoring time, along with a *p* × 1 covariate vector *z*. Let *S*(*t*|*z*) be the survival function of *T* in presence of the covariate vector *z* which can be defined as

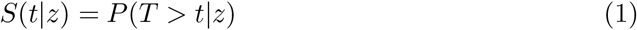

We suppose that some patients will not experience death due COVID-19, even if they are observed infinitely long and our aim is to estimate those fraction of patients. In cure model, we can define the latent variable *Y* as the indicator event as which takes the value 1, if the individual belongs to uncured group (if it experience the event) and 0 otherwise. Now the lifetime *T* can be decomposed as *T* = *YT*\* + (1 − *Y*)∞, where *T*\* is the lifetime of susceptible (uncured) individuals. We can note that the variable *Y* denote the true event status and the variable *δ* denote the observed failure status and essentially, {*δ_i_* = 1, *i* = 1, 2,.., *n*} ⊂ {*Y_i_* = 1, *i* = 1, 2,.., *n*}.

Now, survivor function given in Equation (1) can be written as

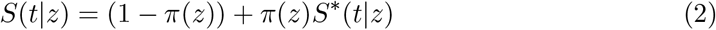

where *S*^*^ (*t*|*z*) is defined as *S*^*^(*t*|*z*) = *P*(*T* > *t*|*z*), which is a proper survivor function. The researchers usually estimate *π*(*z*) by modelling it as logistic distribution given by (Farewell, 1982)

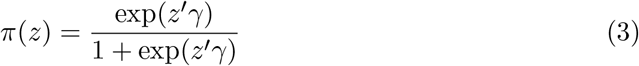

where *γ* is a collection of parameter vectors. In presence of long term survivors, the survivor function *S*(*t*|*z*) is such that lim*_t_*_→_*S*(*t*|*z*) > 0, and this limiting value given by 1 − *π*(*z*), corresponds to the proportion of cured subjects, known as cure rate.

To assess the impact of covariate values *z* on the survivor function of uncured individuals, we can model the survivor function *S*^*^(*t*|*z*). In this paper, we use Cox PH model which is the most popular regression model used in medical research, to model survivor function in presence of covariates. In Cox PH model, the survivor function *S*^*^(*t*|*z*) can be written as

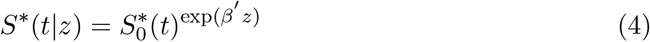

where 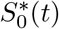 is the baseline survivor function, which is common for all individuals and *β* is the *p* × 1 vector of regression parameters. Now Eq. (4) can be used to model Eq. (2) and the resulting model can be termed as Proportional Hazards Mixture Cure (PHMC) model.

To get a basic information of the presence of cure fraction in a data set, we can use the Kaplan Meier (KM) estimator (Kaplan and Meier, 1958) of baseline survivor function which is independent of covariate vector *z*. The Kaplan Meier or Product Limit estimator at a time *t* is given by

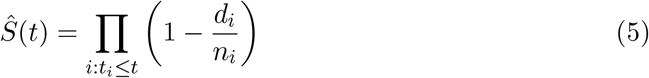

where *d_i_* is the number of events and *n_i_* is the number of individuals at risk at time *t_i_*.

In a perfect situation where all individuals have an observed lifetime we have *S*(*τ*) = 0, where *τ* is the largest observed lifetime. But when cured or long term survivors are presented in the data, obviously *S*(*τ*) > 0 and 1 − *S*(*τ*) will give a rough estimate of the censoring percentage in the data set, which may also include cured proportion. Hence, a high value of *S*(*τ*) gives the evidence of the presence of long term survivors (Maller and Zhao, 1996).

### 2.2. Inference Procedures

Our aim is to estimate the baseline survivor function 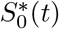 (different from the KM estimator), the vector of regression parameters *β* and the cure fraction *π*(*z*), simultaneously from the given data. To estimate the parameters under a PH model, Peng and Dear (2000) and Sy and Taylor(2000) proposed an partial likelihood method, where we can estimate *β* with out specifying the baseline survivor function 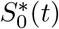. Let Φ = (*T*, *δ*, *z*) denote the observed data.

Expectation-Maximisation (EM) algorithm can be used to estimate the parameters of interest in PHMC model. Following the notations in Section 2.1, given **y** = (*y*_1_, *y*_2_, …,*y_n_*) and Φ, the complete likelihood can be written as

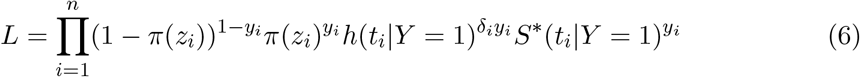

where *h*(.) is the hazard rate function corresponding to *S*^*^(.). The logarithm of the complete likelihood *l* can be written as *l* = *l*_1_ + *l*_2_, where

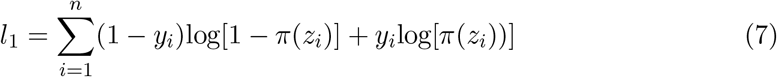

and

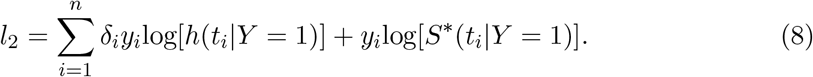

The conditional expectation of the complete log likelihood with respect to *y_i_* given Φ can be calculated using the E-step of EM algorithm along with the estimates of *β* and 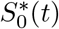. Since Eq. (7) and Eq. (8) are linear functions of we need only the conditional expectation of *y_i_* to perform this computation. Let us denote 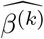 and 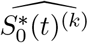 as the estimates of *β* and 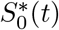 obtained in *k* th iteration. Now the conditional expectation of *y_i_* given 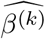 and 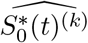 can be written as

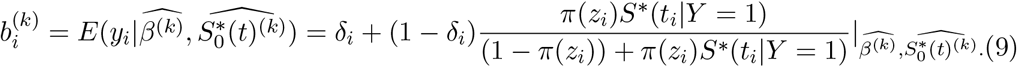

We can see that 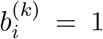 if *δ_i_* = 1 and *δ_i_* = 0, it will be the conditional probability that the *i*th individual remaining uncured. Since 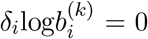 and 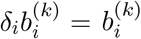, the expectations of Eq. (7) and Eq. (8) can be written as

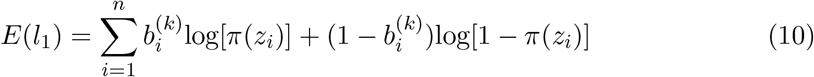

and

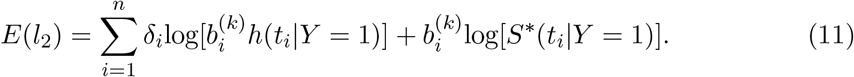

The Maximisation step (M-step) in EM algorithm can be used to maximize Eq. (10) and Eq. (11).

To estimate the parameters under a PH model, we can employ the methods given in Peng and Dear (2000). The log likelihood function under PHMC model can be constructed similar to the usual PH model, with an additional offset variable 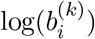. Now the estimate of survivor function using E-step in EM algorithm. Let *t*_(1)_, *t*_(2)_, …*t*_(_*_p_*_)_ denote the distinct uncensored failure times, 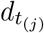 denote the number of events and 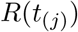 denote the risk set at at time *t*_(_*_j_*_)_. Now the Breslow type estimator for 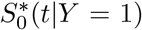 is given by

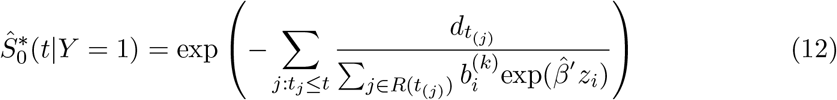

where 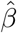 is the estimate of *β*, obtained in the previous step. Now we can estimate the cure probability *π*(*z*) using Eq. (3) given in section 2.1.

## 3. Data and Results

In this Section, we analyse the data on COVID-19 patients in India, using statistical methods explained in Section 2. Since the response variable is time (number of days in hospital) in this study, to employ lifetime data models we need information on the ‘date of admission to hospital’ and ‘status change date’. The available information in the file ‘patient raw data’ from the site https://api.cvoid19india.org as on May 19, 2020 is used to carry out the analysis. From the raw data, we can see that 10.83% of lifetimes are the actually observed lifetimes (current status of the patient is death) and the remaining 89.17% of lifetimes are censored (current status of the patient is deceased/hospitalized).

We plot a Kaplan Meier curve using the Eq. (5) given in Section 2.1, to get a basic information about the presence of long term survivors in the data. The minimum value of survival probability estimated is 0.874 with a standard error 0.0114. The plot of KM curve is given in Figure 1. From the plot it is evident that a large fraction of patients will be long term survivors in this set.

We analyse the above data to estimate the cure fraction in presence of covariates gender and age. Separate analysis is done for both covariates, since for some patients information on gender and for some others information on age is missing. First we estimate the cure fraction considering the covariate gender. In the accessible data, 24.6% are females and remaining 73.4% are males. We denote females by 0 and males by 1 in the analysis. The regression parameter is estimated as 0.386. Since the parameter value is greater than 0 it implies that females will have greater survival probability than males.

**Figure 1.**
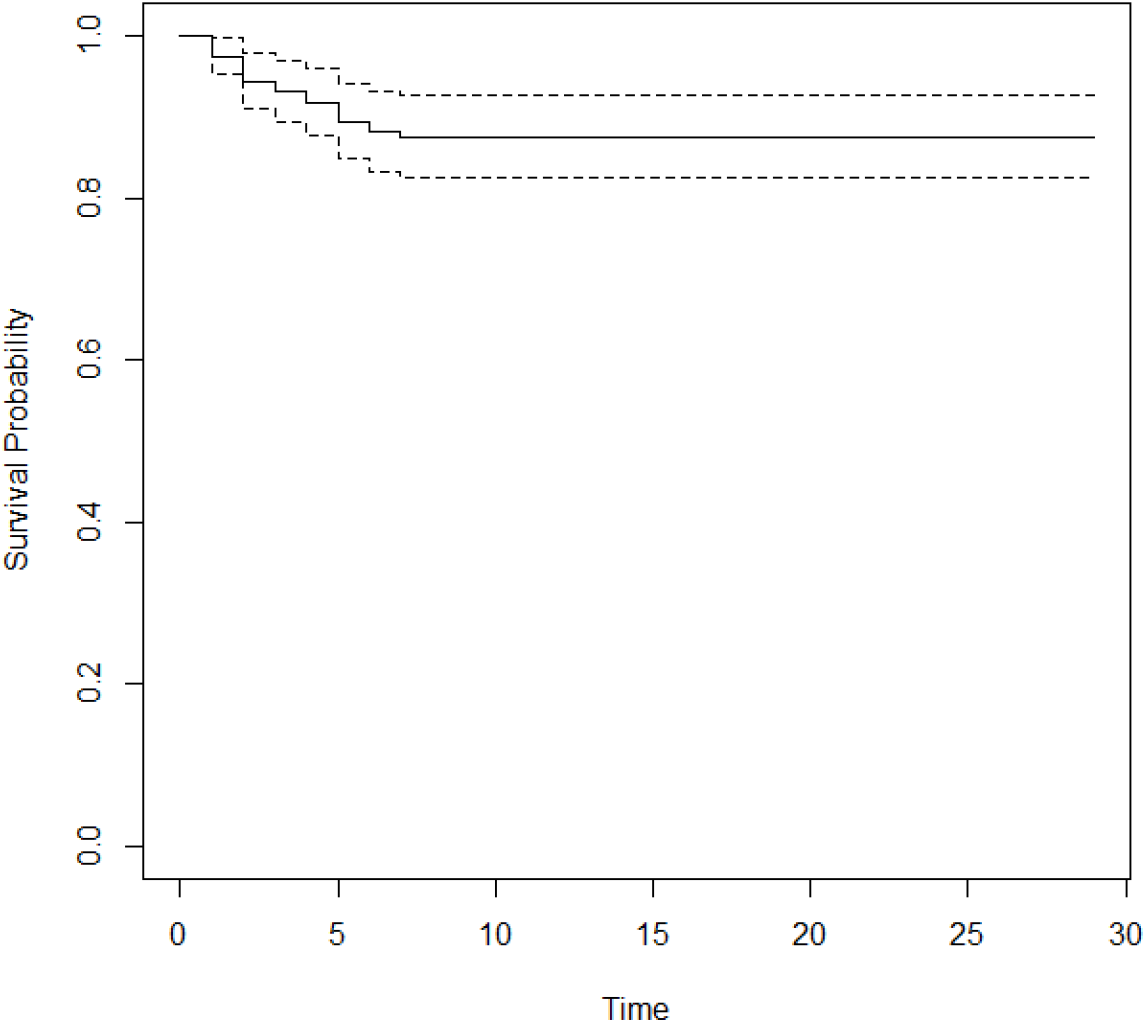
KM plot of COVID-19 patients in India

The cure fraction of the data is estimated as 0.8840. Survival probability predicted using this model is plotted separately for males and females. The plot of the same is given in Figure 2.

In the above figure solid line denotes the survival probability of females and dotted line denote the same of males. We can see that females have a greater survival probability than males; a fact reported by other researchers also. We can also note that the survival probability for females become constant by day 8 and the value is 0.88. In case of males the constancy in survival probability is achieved by day 7 and the value is 0.76.

**Figure 2.**
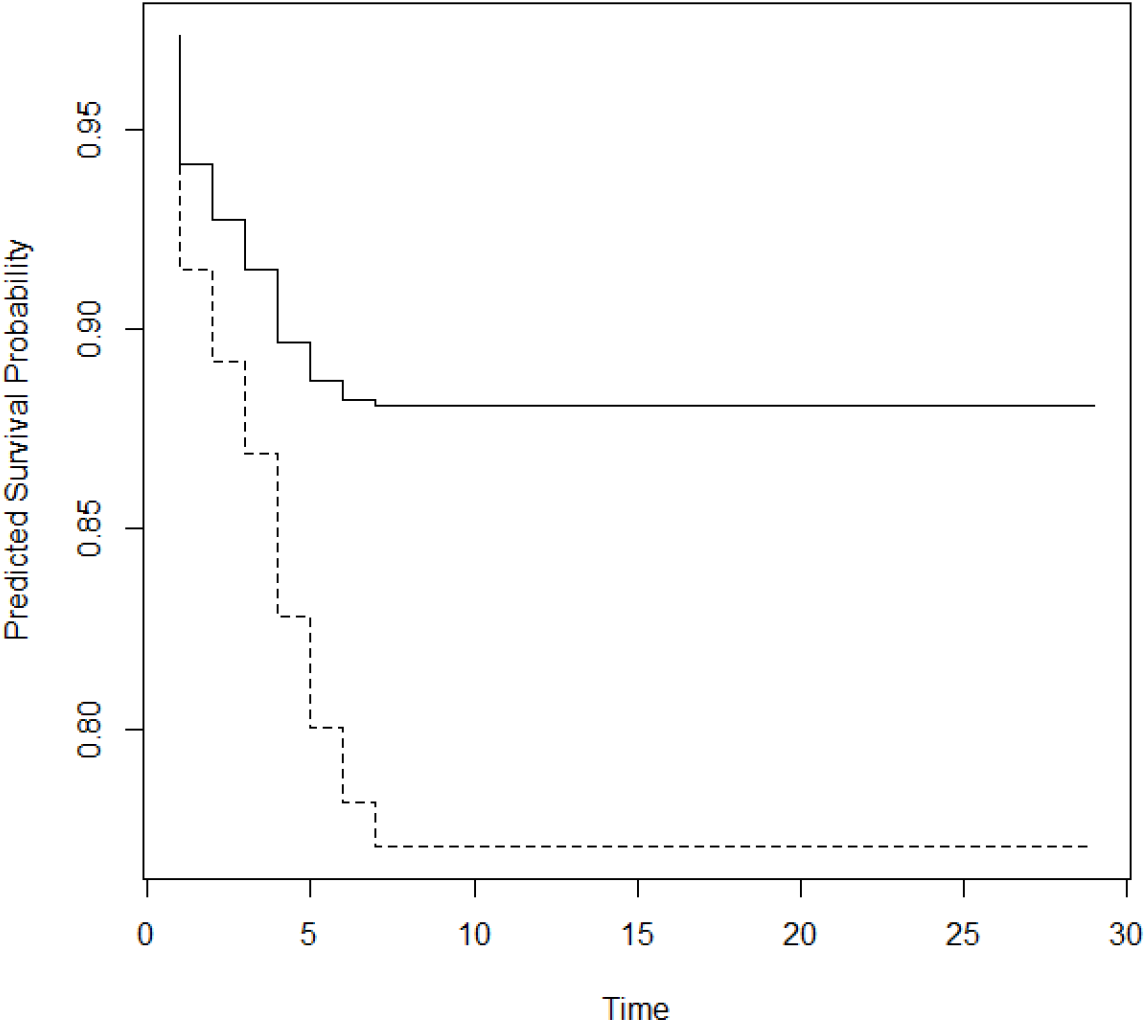
Predicted survival probability of males and females

We now consider the covariate age to estimate cure fraction. The minimum recorded age is 1 and maximum is 96. The regression parameter is estimated as 0.2381, which means that the hazard ratio will be greater than 1. Hence age can be treated as a ‘bad prognostic factor’ which tells us that as age increases hazard will increase. With respect to age, cure fraction is estimated to be 0.9313. We plot the survival probability curve for patients by dividing them in to two groups with respect to age; below 60 years and equal to or above 60 years. The value 60 is chosen to determine, how much the survival pattern is different for senior citizens and others.

In the above figure solid line represents the survival probability of patients below 60 years and dotted line represents the survival probability of patients above 60 years. We can see that there is significant difference in pattern of survival probabilities for senior people and others. For aged people, survival probability became constant after 7 days and it is only 0.5, while for others survival probability attains stability by day 7 and the value is 0.98. This shows that almost all patients of age below 60 are recovering from COVID-19, while nearly half of the aged people are surviving the disease.

**Figure 3.**
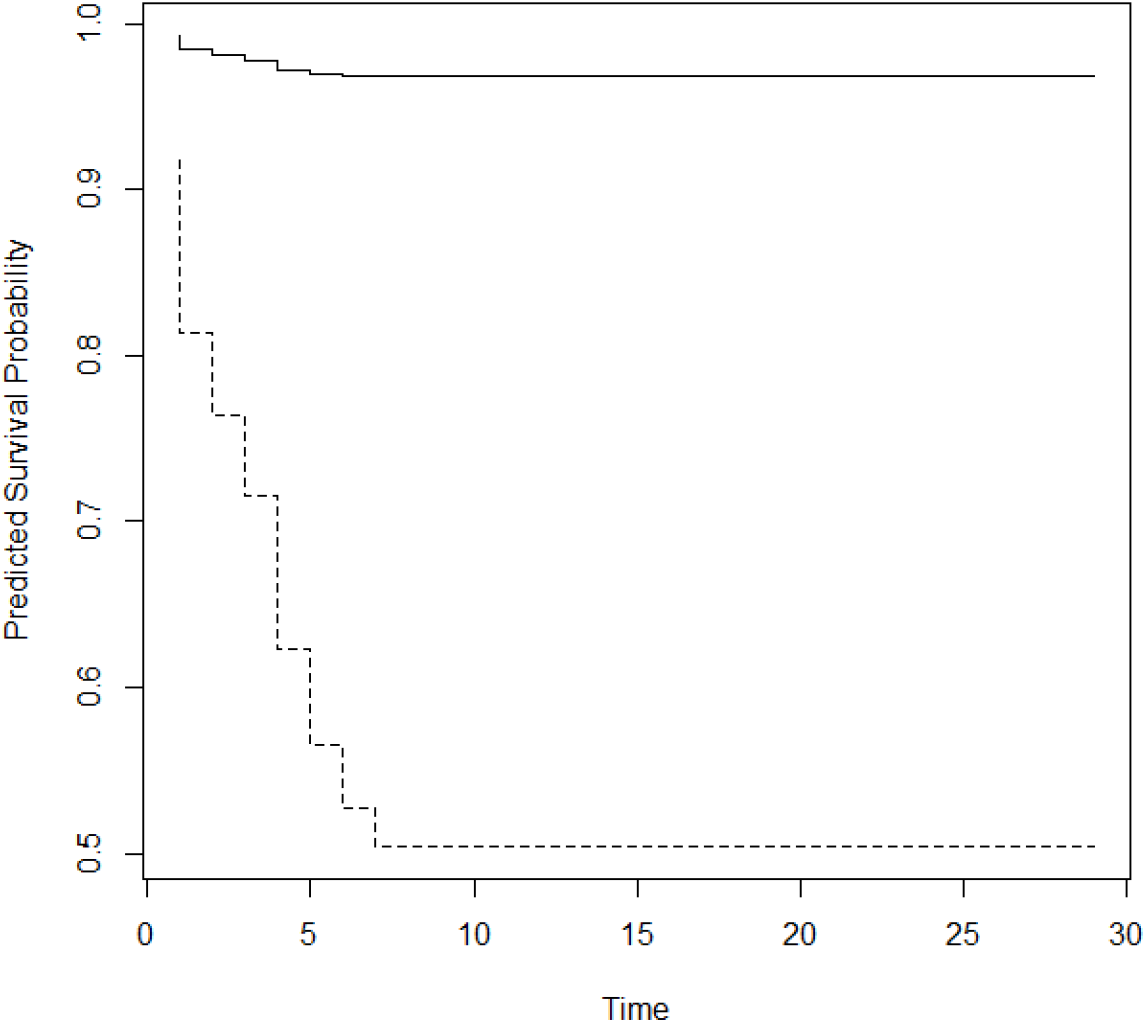
Predicted survival probability of patients above and below 60 years

## 4. Concluding Remarks

We analyse the data on COVID-19 patients in India using statistical methods in lifetime data analysis. All the reported statistical studies on COVID-19 data from various nations use compartmental models in epidemiology. We carry out an explanatory data analysis in the new perspective and estimated the fraction of long term survivors in the data in presence of comorbidities age and gender. It is shown that female patients have a greater chance of survival than male patients. When the younger population has survival probability more than 0.95, for aged population it is around 0.5 only.

A COVID-19 patient entering in the study, may be identified with the disease (or symptoms) at an earlier time, which may unknown. This possibility of partial information on lifetime leads to the data with left censoring. The study can be extended to incorporate left censored individuals also. The grouped data models, where the lifetime is grouped into several non-overlapping groups can also be done in this context. Studies in these directions will be reported elsewhere. Also, it will be worthwhile to analyse the data on COVID-19 patients in presence of information of the current health condition of the patient, since patients with cardiac problems and diabetes may have a greater hazard rate.

## Data Availability

Data available in websites are used in this study.

https://www.worldometers.info/coronavirus/worldwide-graphs

https://www.covid19india.org

https://api.covid19india.org

## Acknowledgements

Sreedevi E. P. would like to thank Kerala State Council for Science Technology and Environment, Kerala, India for the financial support provided to carry out this research work.

The data available on the following websites are used in this study.

https://www.worldometers.info/coronavirus/worldwide-graphs

https://www.covid19india.org

https://api.covid19india.org

